# Measurable Residual *IDH1* before Allogeneic Transplant for Acute Myeloid Leukemia

**DOI:** 10.1101/2023.07.28.23293166

**Authors:** Gege Gui, Laura W. Dillon, Niveditha Ravindra, Pranay S. Hegde, Georgia Andrew, Devdeep Mukherjee, Zoë Wong, Jeffery Auletta, Firas El Chaer, Evan Chen, Yi-Bin Chen, Adam Corner, Steven M. Devine, Sunil Iyer, Antonio Martin Jimenez Jimenez, Marcos J.G. De Lima, Mark R. Litzow, Partow Kebriaei, Stephen R Spellman, Scott L. Zeger, Kristin M. Page, Christopher S. Hourigan

## Abstract

Measurable residual disease (MRD) in adults with acute myeloid leukemia (AML) in complete remission is an important prognostic marker, but detection methodology requires optimization. The persistence of mutated *NPM1* or *FLT3*-ITD in the blood of adult patients with AML in first complete remission (CR1) prior to allogeneic hematopoetic cell transplant (alloHCT) has been established as associated with increased relapse and death after transplant. The prognostic implications of persistence of other common AML-associated mutations, such as *IDH1*, at this treatment landmark however remains incompletely defined. We performed testing for residual *IDH1* variants (*IDH1*m) in pre-transplant CR1 blood of 148 adult patients undergoing alloHCT for *IDH1*-mutated AML at a CIBMTR site between 2013-2019. No post-transplant differences were observed between those testing *IDH1*m positive (n=53, 36%) and negative pre-transplant (overall survival: p = 0.4; relapse: p = 0.5). For patients with *IDH1* mutated AML co-mutated with *NPM1* and/or *FLT3*-ITD, only detection of persistent mutated *NPM1* and/or *FLT3*-ITD was associated with significantly higher rates of relapse (p = 0.01). These data, from the largest study to date, do not support the detection of *IDH1* mutation in CR1 blood prior to alloHCT as evidence of AML MRD or increased post-transplant relapse risk.

## Introduction

Detection of measurable residual disease (MRD) prior to allogeneic hematopoetic cell transplant (alloHCT) in patients with acute myeloid leukemia (AML) in remission is associated with increased relapse and and worse survival after transplant^1-3^. It has been recently reported that detection by ultra-sensitive next generation sequencing (NGS) of mutated *NPM1* or *FLT3* internal tandem duplication (*FLT3*-ITD) persistence in first complete remission (CR1) blood is strongly associated with increased relapse and death compared to testing negative^4^. The genomic etiology of AML is heterogenous and well-described^5^ but the prognostic implication of detecting persistence of all common AML-associated mutations in cytomorphological complete remission remains incompletely defined. Variants in isocitrate dehydrogenase (*IDH*) genes are observed in around 20% of patients diagnosed with AML (7% for *IDH1*), with clinical outcomes reported to differ due to co-occurance of other mutations, *IDH* subtypes, treatment strategies, and patient factors^6-10^. Targeted therapy for *IDH1*-mutated AML is available, and an association between residual *IDH1* mutations and post-transplant clinical outcomes may help provide evidence regarding post-alloHCT maintenance approaches^11-14^. Studies have examined the relationship between residual *IDH1* in CR and subsequent clinical outcomes^15,16^, but definitive nation-level standardized evidence to inform decision making for the clinical utility of *IDH1* as a target for AML MRD testing was not previously available.

## Methods

Adult patients (18 years or older) undergoing first alloHCT for *IDH1* mutated AML in CR1 at a Center for International Blood and Marrow Transplant Research (CIBMTR) site in the United States between 2013-2019 who participated in the CIBMTR data database (NCT01166009) or repository (NCT04920474) protocols, had an available remission blood samples collected within 100 days before alloHCT, and follow-up clinical data including relapse and survival were included in this study. Genomic DNA was extracted from remission blood and sequenced as described previously^4^. The day of transplant was considered as day 0. Median follow-up time was calculated for censored patients. Overall survival (OS) was estimated by Kaplan-Meier (log rank tests) and Cox proportional hazards models (forward selection); cumulative incidence of relapse was examined by Fine-Gray regression models with non-relapse mortality as a competing risk. Additional details are provided in the supplement.

## Results and Discussion

148 patients with *IDH1* mutated AML and undergoing alloHCT in CR1 at a CIBMTR reporting site between 2013 and 2019 were included in the study, with a median follow-up time of 24 months. Relapses were reported in 37 patients (25%), the majority occurring within 1 year after alloHCT (n = 28, 76%; supplemental Figure 1). Pre-transplant remission flow cytometry results were reported to the CIBMTR by the transplant centers for 97% of the patients (n = 144) with a positivity rate of 8% (n = 11). Although there were no statistically significant differences in clinical outcomes when comparing the positive and negative groups (OS: p = 0.3; relapse: p = 0.07; Figure 1A, supplemental Figure 2), patients testing positive by flow cytometry pre-transplant had a trend of higher cumulative incidence of relapse 6 months after alloHCT.

**Figure 1.**
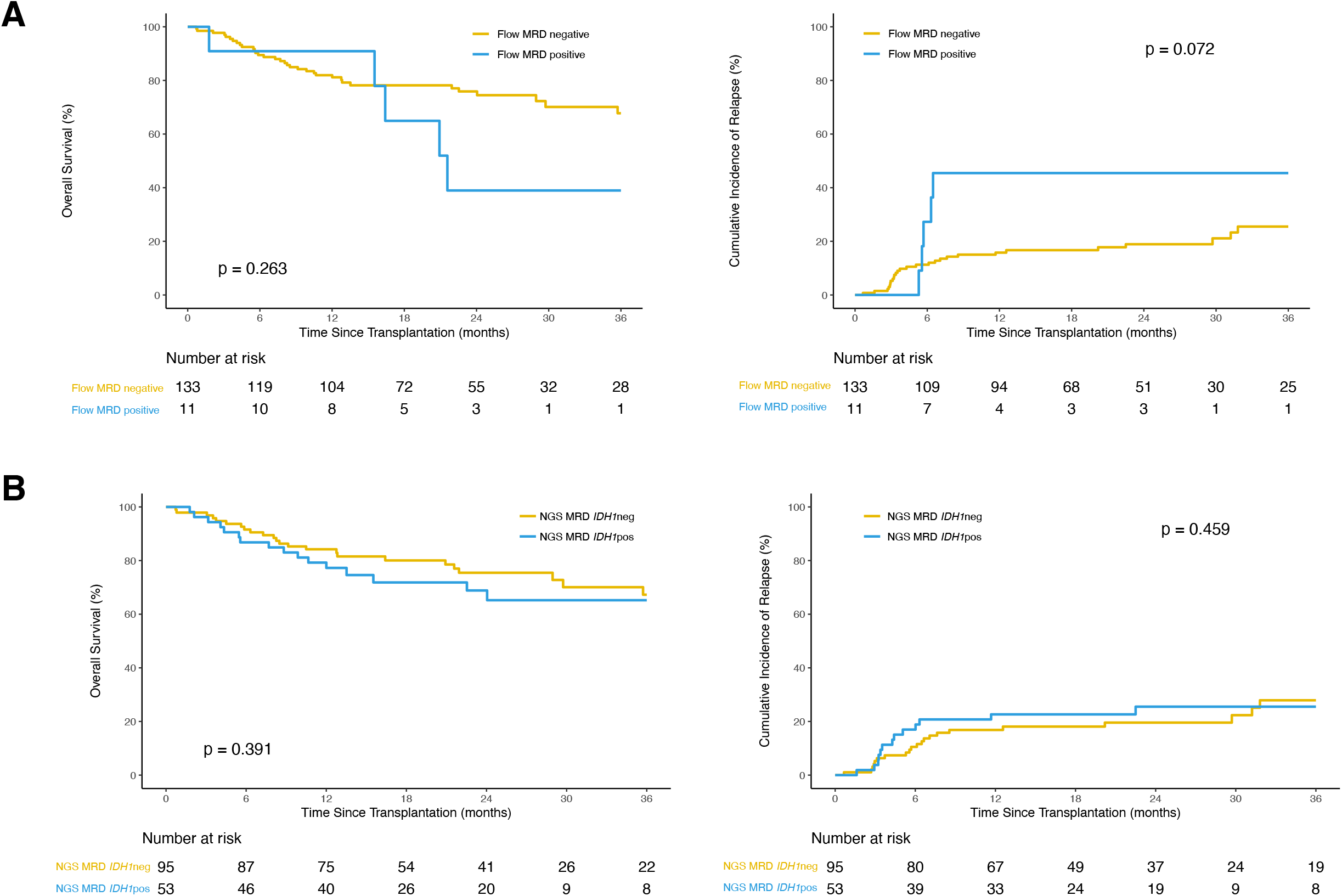
Clinical outcomes for *IDH1*-mutated AML patients after allogeneic hematopoetic cell transplant based on pre-transplant measurable residual disease (MRD) status. **(A)** Overall survival (OS; left) and cumulative incidence of relapse (CIR; with non-relapse mortality as competing risk; right) for patients with available flow cytometry MRD reported by sites (n = 144) grouped by site-reported flow MRD positive (blue) or negative (yellow). **(B)** OS (left) and CIR (with non-relapse mortality as competing risk; right) in patients with *IDH1* mutated AML (n=148) based on the presence (blue) or absence (yellow) of detectable residual *IDH1* variants in the blood pre-transplant during CR1.

NGS MRD analysis identified residual *IDH1* mutations in the pretransplant remission blood of 53 (36%) patients at a median variant allele fraction (VAF) of 1.5% (range 0.09-48.5%), with a 100% validation rate by digital droplet PCR for the variants with an orthogonal assay available (supplemental Table 2, supplemental Figure 3). Logistic regression indicated that *IDH1* NGS-MRD positivity was more likely in patients older than 40yrs (OR: 1.02 with every 1-year increase after 40, p = 0.04). No statistically significant difference between the NGS-MRD *IDH1* positive and negative groups in clinical outcomes was detected (Figure 1B, supplemental Figure 4A), which remained true after further subgroup analyses by age (supplemental Figure 4B) or when stratifying by high (≥ 2.5%) or low VAF groups (<2.5%, supplemental Figure 4C).

Patients with *IDH1* mutated AML also often have mutations in *NPM1* and/or *FLT3*-ITD^5^. Our previous study focusing on patients with *NPM1* and/or *FLT3*-ITD mutated AML showed that patients with either of these mutations detected in CR1 prior to alloHCT had increased relapse and lower OS^4^. Within this cohort of *IDH1* mutated patients, we had information on the baseline mutational status of *NPM1* and *FLT3*-ITD, allowing for investigation into the impact of these co-mutations. A subset of 69 patients (47%) were co-mutated for *NPM1* and/or *FLT3*-ITD at baseline, and those with residual *NPM1* and/or *FLT3*-ITD mutations had higher rates of relapse rate compared to those with *IDH1* only or NGS-MRD negative groups (p = 0.01, 43% vs 11% or 15% at 2yr, Figure 2A, supplemental Figure 5A). The cohort excluding patients with baseline mutations *NPM1* and/or *FLT3*-ITD (n = 79, 53%) did not show any significant clinical outcome differences based on *IDH1* persistence (Figure 2B, supplemental Figure 5B).

**Figure 2.**
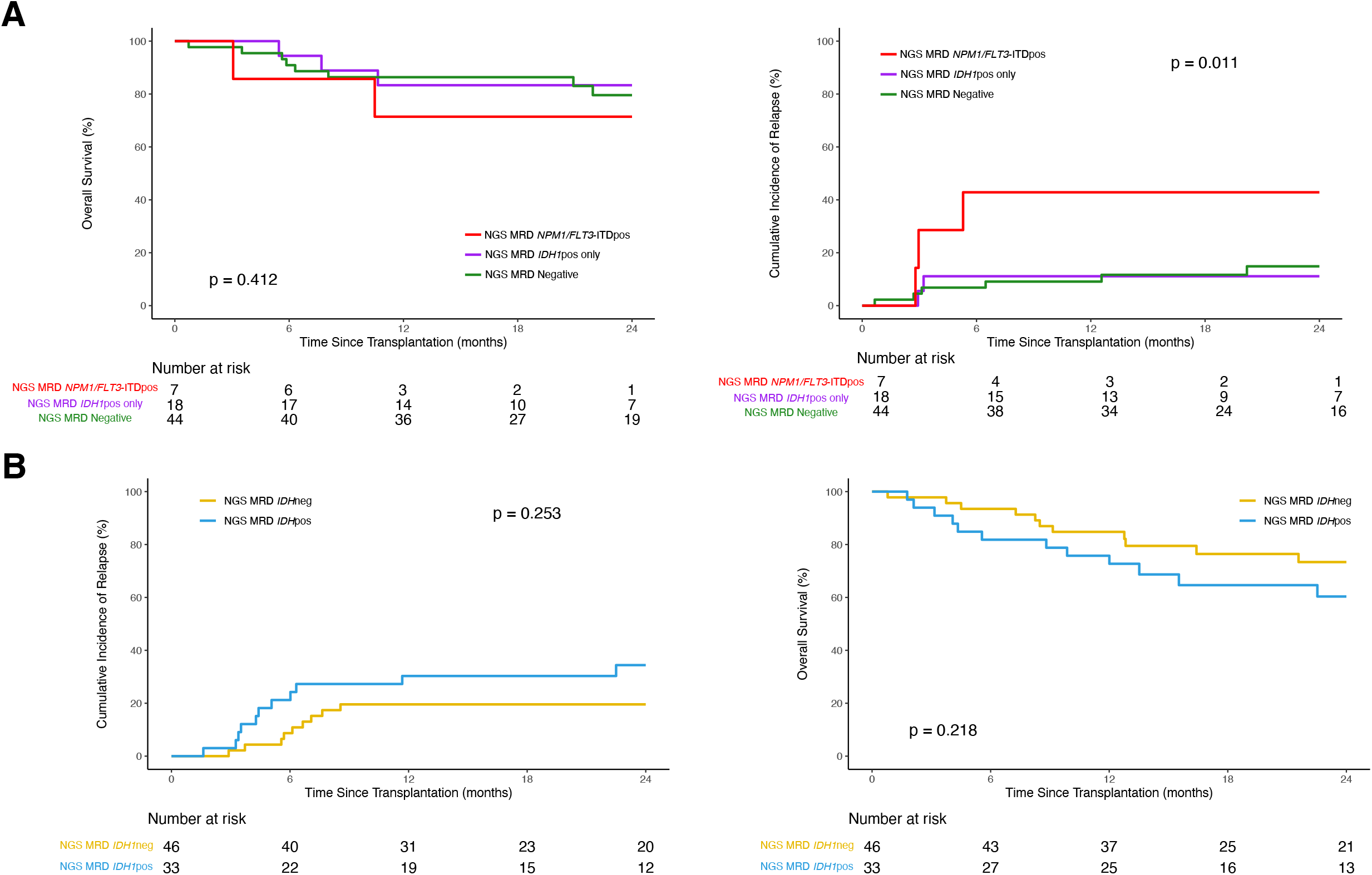
Clinical outcomes for *IDH1*-mutated AML patients after allogeneic hematopoetic cell transplant based on pre-transplant measurable residual disease (MRD) status stratified by co-mutated *NPM1* and/or *FLT3*-ITD at baseline. **(A)** Overall survival (OS; left) and cumulative incidence of relapse (CIR; with non-relapse mortality as competing risk; right) in patients with *IDH1* mutated AML co-mutated with *NPM1* and/or *FLT3*-ITD (n = 69). Patients are stratified based on the presence of residual *NPM1* and/or *FLT3*-ITD variants regardless of residual *IDH1* (red, NGS-MRD *NPM1*/*FLT3*-ITDpos); the presence of residual *IDH1* variants in the absence of residual *NPM1*/*FLT3*-ITD (purple, NGS-MRD *IDH1*pos); and the absence of residual *IDH1, NPM1*, or *FLT3*-ITD variants (green, NGS-MRDneg). **(B)** OS (left) and CIR (with non-relapse mortality as competing risk; right) in patients with *IDH1* mutated AML without baseline *NPM1* and/or *FLT3*-ITD variants (n = 79). Patients are stratified based on the presence (blue) or absence (yellow) of detectable residual *IDH1* variants in the blood pre-transplant during CR1.

We also examined the impact of the conditioning intensity regimens used for alloHCT in these patients and residual *IDH1* variants and saw no association with clinical outcomes (supplemental Figure 6). However, since assignment to conditioning regimen was not randomly assigned, there were more older patients (age > 60yrs) receiving reduced intensity (RIC)/non-myeloablative conditioning (NMA) compared with younger patients (71% vs. 21%; chi-square test: p < 0.001). Additionally, due to small sample size of the subgroups, there were insufficient number of patients to properly examine the interaction between age, conditioning intensity and residual *IDH1* mutations.

The goal of this study was to determine for AML patients the association between pre-alloHCT persistent *IDH1* mutations and post-transplant clinical outcomes. We show that the detection of persistent *IDH1* mutations in the blood of adult patients with AML in CR1 prior to first alloHCT is common and not associated with increased relapse or death after transplant compared to those testing negative. For patients with *IDH1* mutated AML co-mutated with *NPM1* and/or *FLT3*-ITD, detection of persistent *NPM1* and/or *FLT3*-ITD was associated with higher rates of relapse. These data, from the largest study to date, do not support the detection of isolated *IDH1* mutation in CR1 blood prior to alloHCT as evidence of AML MRD or increased post-transplant risk.

## Data Availability

Raw FASTQ files are the NCBI Sequencing Reads Archive (SRA) (Accession: PRJNA834073 and PRJNA997373).

## Acknowledgement

This work was supported by the Intramural Research Program of the National Heart, Lung, and Blood Institute, the National Institutes of Health Director’s Challenge Innovation Award, and the Foundation of the NIH AML MRD Biomarkers Consortium.

Sequencing was performed in the NHLBI Intramural DNA Sequencing and Genomics Core.

The CIBMTR is supported primarily by Public Health Service U24CA076518 from the National Cancer Institute (NCI), the National Heart, Lung and Blood Institute (NHLBI) and the National Institute of Allergy and Infectious Diseases (NIAID); U24HL138660 and U24HL157560 from NHLBI and NCI; U24CA233032 from the NCI; OT3HL147741 and U01HL128568 from the NHLBI; HHSH250201700005C, HHSH250201700006C, and HHSH250201700007C from the Health Resources and Services Administration (HRSA); and N00014-20-1-2832 and N00014-21-1-2954 from the Office of Naval Research.

The views expressed in this article do not reflect the official policy or position of the National Institute of Health, the Department of the Navy, the Department of Defense, Health Resources and Services Administration (HRSA) or any other agency of the U.S. Government.

## Conflicts of Interest Statements

Hourigan: The National Heart, Lung, and Blood Institute receives research funding for the laboratory of Dr. Hourigan from the Foundation of the NIH AML MRD Biomarkers Consortium.

Auletta: Advisory Committee: AscellaHealth and Takeda

El Chaer: Consultant: SPD Oncology, Amgen, Association of Community Cancer Centers; Clinical Trial Grant Support (PI) to the University of Virginia: Amgen, BMS, Celgene, SPD Oncology, Sanofi, Bristol Myers Squibb, FibroGen, PharmaEssentia, BioSight, MEI Pharma, Novartis, Arog pharmaceuticals; Travel grant: DAVA Oncology

E Chen: Consultant: Rigel Pharmaceuticals and AbbVie

Corner: Employment: Bio-Rad Laboratories

Jimenez Jimenez: Funding: Abbvie

De Lima: Advisory Board: Pfizer, Bristol Myers Squibb; Data Safety Monitoring Board: Novartis, Abbvie; Research Funding: Miltenyi Biotec

Kebriaei: Consultant: Pfizer, Jazz Pharmaceuticals

## Notes

### Funding Statement

This work was supported in part by the Intramural Research Program of the National Heart, Lung, and Blood Institute and by the National Institutes of Health Director's Challenge Innovation Award, and the Foundation of the NIH AML MRD Biomarkers Consortium.. The CIBMTR is supported primarily by Public Health Service U24CA076518 from the National Cancer Institute (NCI), the National Heart, Lung and Blood Institute (NHLBI) and the National Institute of Allergy and Infectious Diseases (NIAID); U24HL138660 and U24HL157560 from NHLBI and NCI; U24CA233032 from the NCI; OT3HL147741 and U01HL128568 from the NHLBI; HHSH250201700005C, HHSH250201700006C, and HHSH250201700007C from the Health Resources and Services Administration (HRSA); and N00014-20-1-2832 and N00014-21-1-2954 from the Office of Naval Research.

### Author Declarations

Patients provided written informed consent to participate in the Center for International Blood and Marrow Transplant Research research protocol, which was approved by the National Marrow Donor Program institutional review board.

