## Supplementary Appendix for "Measurable Residual *IDH1* before Allogeneic Transplant for Acute Myeloid Leukemia"

**Table of Contents**


| | |
|---|---|
| Supplemental Methods | Page 2 |
| Supplemental Figure 1. Baseline characteristics for *IDH1*-mutated AML patients and the association with clinical outcomes after allogeneic hematopoietic cell transplant. | Page 4 |
| Supplemental Figure 2. Site-reported flow cytometry MRD status for *IDH1*-mutated AML patients and the association with clinical outcomes after allogeneic hematopoietic cell transplant. | Page 5 |
| Supplemental Figure 3. Detection and validation of residual variants in pretransplant blood of *IDH1* mutated AML patients during complete remission. | Page 6 |
| Supplemental Figure 4. NGS MRD status for *IDH1*-mutated AML patients and the association with clinical outcomes after allogeneic hematopoietic cell transplant stratified by age and variant allele fraction. | Page 7 |
| Supplemental Figure 5. NGS MRD status for *IDH1*-mutated AML patients and the association with clinical outcomes after allogeneic hematopoietic cell transplant stratified by baseline mutation groups. | Page 9 |
| Supplemental Figure 6. NGS MRD status for *IDH1*-mutated AML patients and the association with clinical outcomes after allogeneic hematopoietic cell transplant stratified by conditioning intensity. | Page 10 |
| Supplemental Table 1. *IDH1*-mutated AML patient baseline clinical characteristics. | Page 11 |
| Supplemental Table 2. Variants detected by next-generation sequencing in the blood of *IDH1* mutated AML patients prior to transplant conditioning. | Page 12 |
| References | Page 13 |

### Supplemental Methods

#### Patient baseline characteristics

Patient baseline characteristics included age, sex, race, hematopoietic cell transplant specific comorbidity (HCT-CI), Karnofsky performance status (KPS), secondary AML, European LeukemiaNet (ELN) 2017 risk group, baseline genetics before transplant (*IDH1, IDH2, KIT, NPM1, FLT3-TKD, FLT3*-ITD), conditioning regimen, graft type, donor group, anti-thymocyte globulin (ATG) usage, and site-reported flow cytometry MRD status.

#### Targeted next-generation sequencing (NGS)

Genomic DNA was extracted from remission blood and residual variant identified as previously reported.[1] In short, ultrasensitive error-corrected NGS targeting mutational hotspot regions of the *IDH1*, *IDH2*, *KIT*, *NPM1*, and *FLT3* genes was performed utilizing an automated workflow with pre- and post-PCR separation. NGS libraries were sequencing on a NovaSeq 6000 (Illumina) with unique dual indices. Bioinformatic pipelines were used to perform error-corrected variant calling followed by filtering to identify residual disease. Raw FASTQ files are the NCBI Small Reads Archive (SRA) (Accession: PRJNA834073 and PRJNA997373).

#### Digital droplet PCR (ddPCR)

A subset of 51 *IDH1* variants identified by NGS were orthogonally validated using ddPCR on the Bio-Rad QX200 or QX600 system as previously described.[1]

#### Statistical analysis

Logistic regression was implemented for determining the variables related with NGS *IDH1* MRD status. Overall survival and cumulative incidence of relapse were considered as the primary endpoints, and the follow-up time was collected with day of transplant as time 0. Median follow-up time was calculated for censored patients. Kaplan-Meier estimation and log rank tests were used to calculate overall survival and relapse-free survival endpoints. Cox proportional hazards models were fitted, with forward selection or Lasso penalty for variable selection, and the proportional hazards assumptions were validated. Fine-Gray regression models were used to examine the cumulative incidence of relapse with transplant related mortality as a competing risk, and Bayesian information criterion (BIC) was used for model selection. Potential interactions between mutation groups and clinical characteristics were tested.

**Supplemental Figure 1. Baseline characteristics for *IDH1*-mutated AML patients and the association with clinical outcomes after allogeneic hematopoietic cell transplant.** Cumulative incidence of non-relapse mortality (NRM, top left) and relapse (top right), relapse-free survival (RFS, bottom left) and overall survival (OS, bottom right) shown at 36 months based on baseline patient characteristics (A) overall and (B) age group (<60yrs or ≥60yrs). Point estimates at different time points are shown in the table (far right). Overall P values: Gray's test for non-relapse mortality (NRM) and relapse; log-rank test for relapse-free survival (RFS) and overall survival (OS). P values for pointwise estimations at different time points: z-test. Confidence interval, CI; Probability, prob; Month, mo; Year, yr.

A

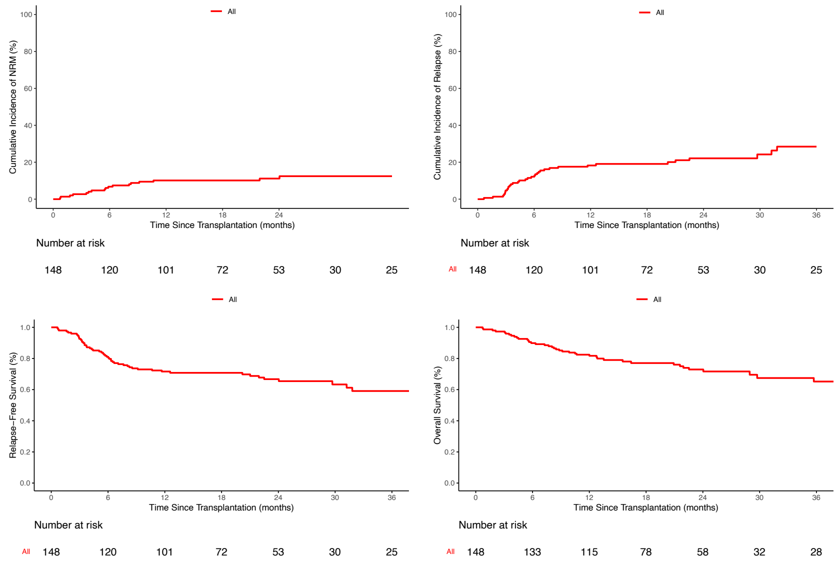


#### All patients

| Outcomes | Prob (95% CI) |
|---|---|
| NRM | Overall n=148 |
| 3 mo | 3(0-5)% |
| 6 mo | 7(3-11)% |
| 1 yr | 10(5-15)% |
| 2 yr | 11(6-16)% |
| 3 yr | 12(7-18)% |
| Relapse | Overall n=148 |
| 3 mo | 5(1-8)% |
| 6 mo | 13(7-18)% |
| 1 yr | 19(13-25)% |
| 2 yr | 22(15-29)% |
| 3 yr | 28(19-37)% |
| RFS | Overall n=148 |
| 3 mo | 93(87-96)% |
| 6 mo | 80(73-86)% |
| 1 yr | 71(63-78)% |
| 2 yr | 67(59-74)% |
| 3 yr | 60(50-69)% |
| OS | Overall n=148 |
| 3 mo | 97(93-99)% |
| 6 mo | 90(84-94)% |
| 1 yr | 82(75-88)% |
| 2 yr | 73(64-80)% |
| 3 yr | 66(55-75)% |

B

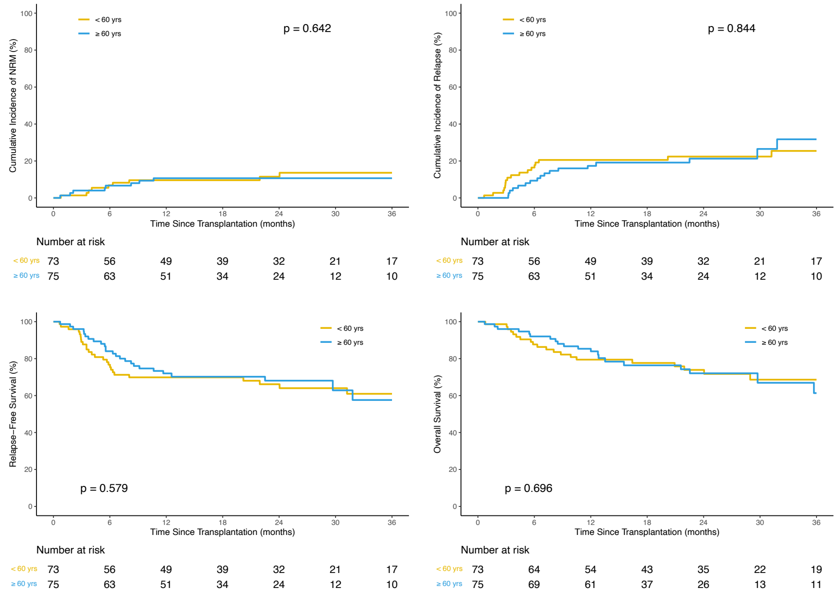


#### Age Group

| Outcomes | Prob (95% CI) | Prob (95% CI) | p-value |
|---|---|---|---|
| NRM | <60yrs=73 | ≥60yrs=75 | 0.642 |
| 3 mo | 1(0-4)% | 4(0-8)% | 0.322 |
| 6 mo | 7(1-13)% | 7(1-12)% | 0.965 |
| 1 yr | 10(3-16)% | 11(4-18)% | 0.829 |
| 2 yr | 11(4-19)% | 11(4-18)% | 0.878 |
| 3 yr | 14(5-22)% | 11(4-18)% | 0.602 |
| Relapse | <60yrs=73 | ≥60yrs=75 | 0.844 |
| 3 mo | 10(3-16)% | 0(0-0)% | 0.006 |
| 6 mo | 16(8-25)% | 9(3-16)% | 0.199 |
| 1 yr | 21(11-30)% | 17(9-26)% | 0.62 |
| 2 yr | 22(13-32)% | 21(11-31)% | 0.874 |
| 3 yr | 25(14-37)% | 32(15-48)% | 0.533 |
| RFS | <60yrs=73 | ≥60yrs=75 | 0.579 |
| 3 mo | 89(79-94)% | 96(88-99)% | 0.106 |
| 6 mo | 77(65-85)% | 84(74-91)% | 0.263 |
| 1 yr | 70(58-79)% | 72(60-81)% | 0.775 |
| 2 yr | 66(54-76)% | 68(56-78)% | 0.808 |
| 3 yr | 61(47-72)% | 58(40-72)% | 0.748 |
| OS | <60yrs=73 | ≥60yrs=75 | 0.696 |
| 3 mo | 99(91-100)% | 96(88-99)% | 0.319 |
| 6 mo | 88(78-93)% | 92(83-96)% | 0.383 |
| 1 yr | 79(68-87)% | 85(75-92)% | 0.347 |
| 2 yr | 74(62-83)% | 72(59-82)% | 0.816 |
| 3 yr | 69(55-79)% | 61(42-76)% | 0.492 |

**Supplemental Figure 2. Site-reported flow cytometry MRD status for *IDH1*-mutated AML patients and the association with clinical outcomes after allogeneic hematopoietic cell transplant.** Cumulative incidence of non-relapse mortality (NRM, top left) and relapse (top right), relapse-free survival (RFS, bottom left) and overall survival (OS, bottom right) shown at 36 months for patients with data available for residual disease by flow cytometry as reported by the treatment site. Point estimates at different time points are shown in the table (far right). Overall P values: Gray's test for non-relapse mortality (NRM) and relapse; log-rank test for relapse-free survival (RFS) and overall survival (OS). P values for pointwise estimations at different time points: z-test. Confidence interval, CI; Probability, prob; Month, mo; Year, yr.

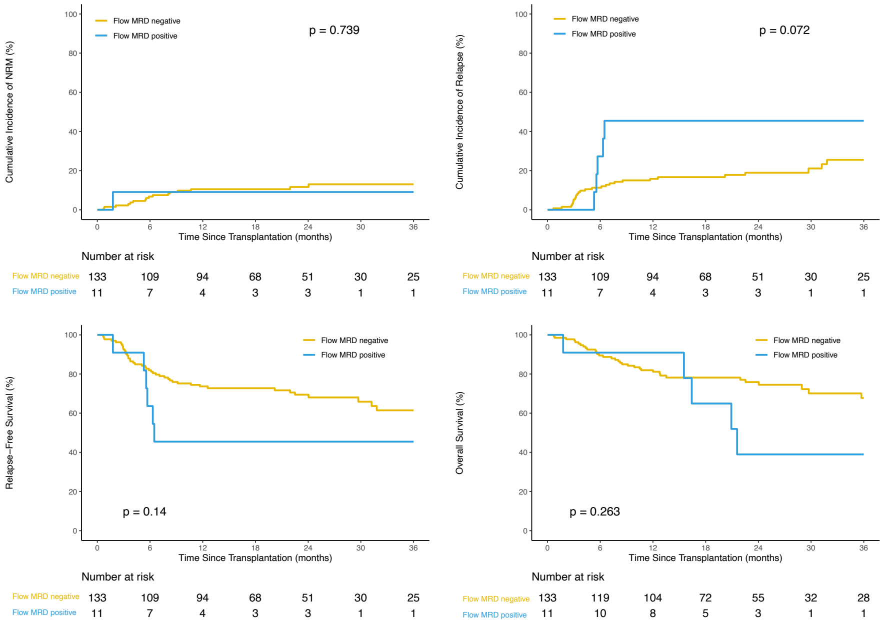


**Site-Reported Flow Cytometry MRD**

| Outcomes | Prob (95% CI) | Prob (95% CI) | p-value |
|---|---|---|---|
| NRM | Flow MRD negative n =133 | Flow MRD Positive n =11 | 0.739 |
| 3 mo | 2(0-5)% | 9(0-27)% | 0.457 |
| 6 mo | 7(2-11)% | 9(0-27)% | 0.804 |
| 1 yr | 11(5-16)% | 9(0-27)% | 0.88 |
| 2 yr | 12(6-17)% | 9(0-27)% | 0.789 |
| 3 yr | 13(7-19)% | 9(0-27)% | 0.682 |
| Relapse | Flow MRD negative n =133 | Flow MRD Positive n =11 | 0.072 |
| 3 mo | 5(1-9)% | 0(0-0)% | 0.007 |
| 6 mo | 11(6-17)% | 27(0-55)% | 0.271 |
| 1 yr | 16(10-22)% | 45(14-77)% | 0.071 |
| 2 yr | 19(12-26)% | 45(14-77)% | 0.108 |
| 3 yr | 26(16-35)% | 45(14-77)% | 0.237 |
| RFS | Flow MRD negative n =133 | Flow MRD Positive n =11 | 0.14 |
| 3 mo | 92(86-96)% | 91(51-99)% | 0.861 |
| 6 mo | 82(74-88)% | 64(30-85)% | 0.218 |
| 1 yr | 74(65-80)% | 45(17-71)% | 0.068 |
| 2 yr | 69(60-77)% | 45(17-71)% | 0.124 |
| 3 yr | 61(50-71)% | 45(17-71)% | 0.315 |
| OS | Flow MRD negative n =133 | Flow MRD Positive n =11 | 0.263 |
| 3 mo | 98(93-99)% | 91(51-99)% | 0.435 |
| 6 mo | 89(83-94)% | 91(51-99)% | 0.874 |
| 1 yr | 82(74-88)% | 91(51-99)% | 0.335 |
| 2 yr | 76(67-83)% | 39(9-69)% | 0.038 |
| 3 yr | 68(56-77)% | 39(9-69)% | 0.113 |

**Supplemental Figure 3. Detection of residual variants in pretransplant blood of *IDH1* mutated AML patients during complete remission.** (A) The total number and (B) variant allele fraction (VAF) of variants per gene as detected by targeted next-generation sequencing (NGS) during remission prior to transplant in the peripheral blood of *IDH1* mutated AML patients. (C) A total of 51 of the 53 detected variants in *IDH1* had a validated assay available for orthogonal validation by digital droplet PCR (ddPCR). The VAF of *IDH1* variants detected by NGS (x-axis) were plotted versus ddPCR (y-axis), with a value of equivalence line displayed as a dotted line. A significant correlation was observed in the VAF as detected by NGS compared to ddPCR with an orthogonal validation rate of 100%. The Pearson correlation coefficient is shown on the inset of the graph.

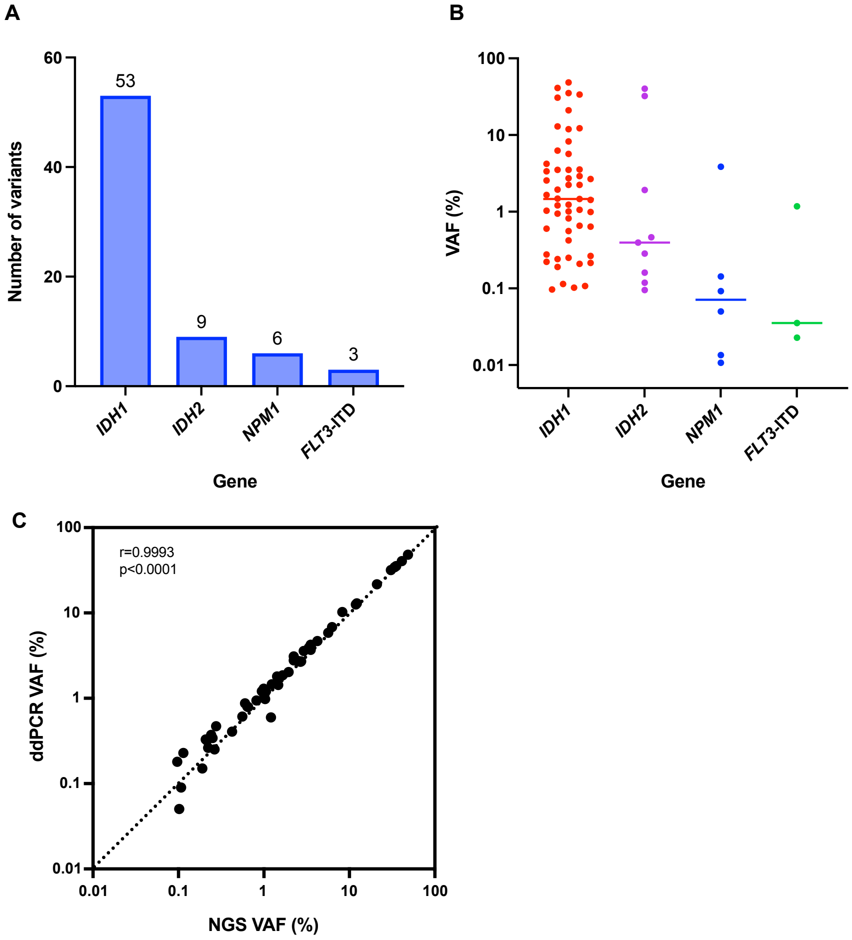

**Supplemental Figure 4. NGS MRD status for *IDH1*-mutated AML patients and the association with clinical outcomes after allogeneic hematopoietic cell transplant stratified by age and variant allele fraction (VAF).** Cumulative incidence of non-relapse mortality (NRM, top left) and relapse (top right), relapse-free survival (RFS, bottom left) and overall survival (OS, bottom right) shown at 36 months based on *IDH1* NGS-MRD status for (A) all patients (B) patients defined by age group (<60yrs or ≥60yrs), and (C) *IDH1* NGS-MRD VAF groups (negative, 0%<VAF<2.5%, VAF≥2.5%). Point estimates at different time points are shown in the table (far right). Overall P values: Gray's test for non-relapse mortality (NRM) and relapse; log-rank test for relapse-free survival (RFS) and overall survival (OS). P values for pointwise estimations at different time points: z-test. Confidence interval, CI; Probability, prob; Month, mo; Year, yr; pos, positive; neg, negative.

**A**

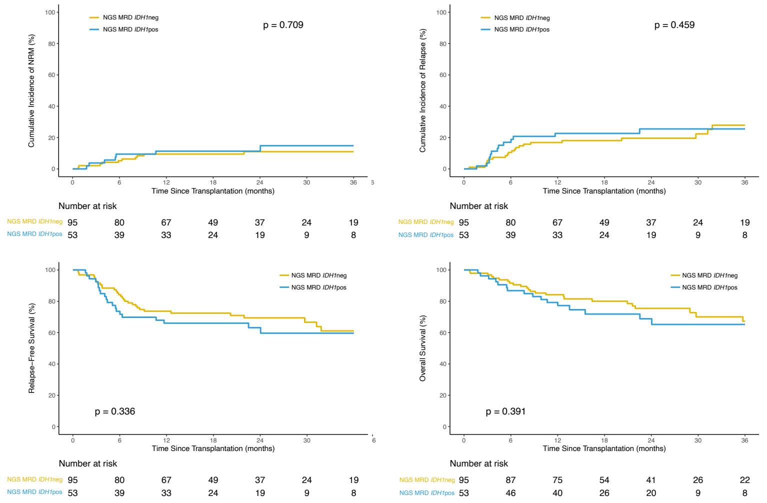


**NGS MRD *IDH1***

| Outcomes | Prob (95% CI) | Prob (95% CI) | p-value |
|---|---|---|---|
| NRM | NGS MRD *IDH1*neg n=95 | NGS MRD *IDH1*pos n=53 | 0.709 |
| 3 mo | 2(0-5)% | 4(0-9)% | 0.582 |
| 6 mo | 5(1-10)% | 9(1-17)% | 0.372 |
| 1 yr | 9(4-15)% | 11(3-20)% | 0.729 |
| 2 yr | 11(4-18)% | 11(3-20)% | 0.956 |
| 3 yr | 11(4-18)% | 15(4-26)% | 0.554 |
| Relapse | NGS MRD *IDH1*neg n=95 | NGS MRD *IDH1*pos n=53 | 0.459 |
| 3 mo | 5(1-10)% | 4(0-9)% | 0.671 |
| 6 mo | 11(4-17)% | 17(7-27)% | 0.29 |
| 1 yr | 17(9-24)% | 23(11-34)% | 0.406 |
| 2 yr | 20(11-28)% | 26(13-38)% | 0.432 |
| 3 yr | 28(16-40)% | 26(13-38)% | 0.783 |
| RFS | NGS MRD *IDH1*neg n=95 | NGS MRD *IDH1*pos n=53 | 0.336 |
| 3 mo | 93(85-96)% | 92(81-97)% | 0.968 |
| 6 mo | 84(75-90)% | 74(60-83)% | 0.136 |
| 1 yr | 74(64-81)% | 66(52-77)% | 0.334 |
| 2 yr | 69(59-78)% | 63(48-75)% | 0.457 |
| 3 yr | 61(48-72)% | 60(44-72)% | 0.882 |
| OS | NGS MRD *IDH1*neg n=95 | NGS MRD *IDH1*pos n=53 | 0.391 |
| 3 mo | 98(92-99)% | 96(86-99)% | 0.579 |
| 6 mo | 92(84-96)% | 87(74-93)% | 0.38 |
| 1 yr | 84(75-90)% | 79(66-88)% | 0.459 |
| 2 yr | 75(65-83)% | 69(53-80)% | 0.43 |
| 3 yr | 67(54-78)% | 65(49-78)% | 0.832 |

**B**

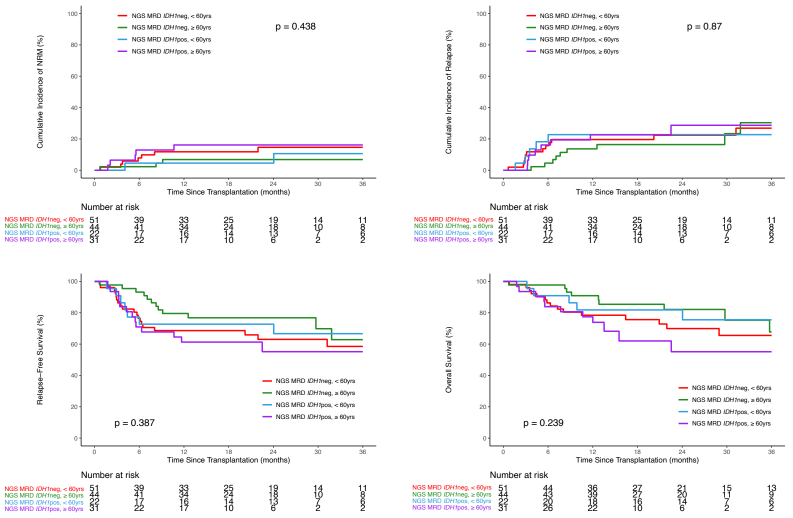


**NGS MRD *IDH1* and Age Group**

| Outcomes | Prob (95% CI) | Prob (95% CI) | Prob (95% CI) | Prob (95% CI) | p-value |
|---|---|---|---|---|---|
| NRM | NGS MRD *IDH1*neg <60yrs: n=51 | NGS MRD *IDH1*neg ≥60yrs: n=44 | NGS MRD *IDH1*pos <60yrs: n=22 | NGS MRD *IDH1*pos ≥60yrs: n=31 | 0.438 |
| 3 mo | 2(0-6)% | 2(0-7)% | 0(0-0)% | 6(0-15)% | 0.254 |
| 6 mo | 8(0-15)% | 2(0-7)% | 5(0-13)% | 13(1-25)% | 0.308 |
| 1 yr | 12(3-21)% | 7(0-14)% | 5(0-13)% | 16(3-29)% | 0.435 |
| 2 yr | 15(4-25)% | 7(0-14)% | 5(0-13)% | 16(3-29)% | 0.317 |
| 3 yr | 15(4-25)% | 7(0-14)% | 11(0-25)% | 16(3-29)% | 0.528 |
| Relapse | NGS MRD *IDH1*neg <60yrs: n=51 | NGS MRD *IDH1*neg ≥60yrs: n=44 | NGS MRD *IDH1*pos <60yrs: n=22 | NGS MRD *IDH1*pos ≥60yrs: n=31 | 0.87 |
| 3 mo | 10(2-18)% | 0(0-0)% | 9(0-21)% | 0(0-0)% | NA |
| 6 mo | 16(6-26)% | 5(0-11)% | 18(2-35)% | 16(3-29)% | 0.116 |
| 1 yr | 20(9-31)% | 14(3-24)% | 23(5-41)% | 23(8-38)% | 0.708 |
| 2 yr | 22(10-34)% | 16(5-28)% | 23(5-41)% | 29(10-47)% | 0.705 |
| 3 yr | 27(13-41)% | 30(10-51)% | 23(5-41)% | 29(10-47)% | 0.95 |
| RFS | NGS MRD *IDH1*neg <60yrs: n=51 | NGS MRD *IDH1*neg ≥60yrs: n=44 | NGS MRD *IDH1*pos <60yrs: n=22 | NGS MRD *IDH1*pos ≥60yrs: n=31 | 0.387 |
| 3 mo | 88(76-95)% | 98(85-100)% | 91(68-98)% | 94(77-98)% | 0.233 |
| 6 mo | 76(62-86)% | 93(80-98)% | 77(54-90)% | 71(52-84)% | 0.015 |
| 1 yr | 69(54-79)% | 80(64-89)% | 73(49-87)% | 61(42-76)% | 0.346 |
| 2 yr | 63(48-75)% | 77(61-87)% | 73(49-87)% | 55(34-72)% | 0.23 |
| 3 yr | 59(42-72)% | 63(39-79)% | 67(42-83)% | 55(34-72)% | 0.86 |
| OS | NGS MRD *IDH1*neg <60yrs: n=51 | NGS MRD *IDH1*neg ≥60yrs: n=44 | NGS MRD *IDH1*pos <60yrs: n=22 | NGS MRD *IDH1*pos ≥60yrs: n=31 | 0.239 |
| 3 mo | 98(87-100)% | 98(85-100)% | 100% | 94(77-98)% | <0.001 |
| 6 mo | 86(73-93)% | 98(85-100)% | 91(68-98)% | 84(66-93)% | 0.048 |
| 1 yr | 78(64-87)% | 91(78-96)% | 82(59-93)% | 77(58-89)% | 0.232 |
| 2 yr | 70(54-81)% | 82(66-91)% | 82(59-93)% | 55(31-74)% | 0.127 |
| 3 yr | 66(48-78)% | 68(43-84)% | 76(50-89)% | 55(31-74)% | 0.586 |

**C**

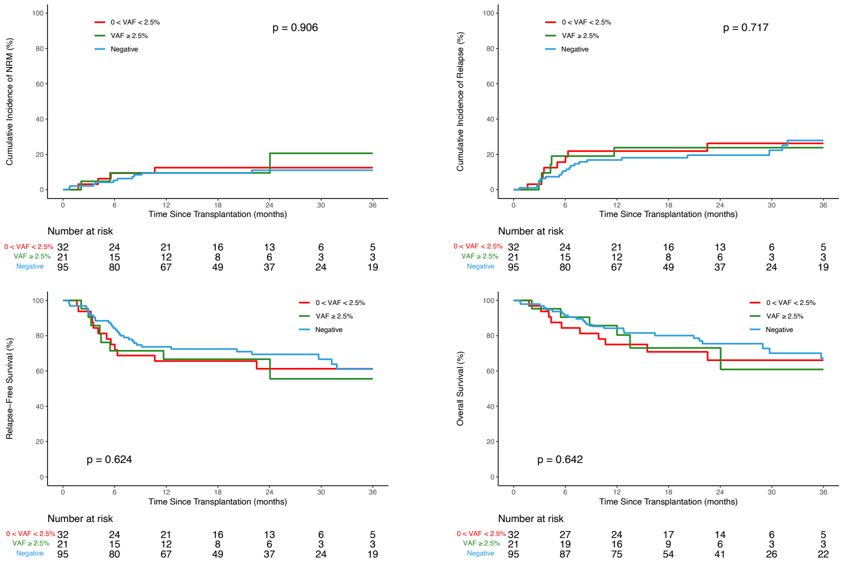


**VAF Groups**

| Outcomes | Prob (95% CI) | Prob (95% CI) | Prob (95% CI) | p-value |
|---|---|---|---|---|
| NRM | 0<VAF<2.5% =32 | VAF ≤ 2.5% =21 | Negative=95 | 0.906 |
| 3 mo | 3(0-9)% | 5(0-14)% | 2(0-5)% | 0.845 |
| 6 mo | 9(0-20)% | 10(0-22)% | 5(1-10)% | 0.676 |
| 1 yr | 12(1-24)% | 10(0-22)% | 9(4-15)% | 0.9 |
| 2 yr | 12(1-24)% | 10(0-22)% | 11(4-18)% | 0.945 |
| 3 yr | 12(1-24)% | 21(0-45)% | 11(4-18)% | 0.756 |
| Relapse | 0<VAF<2.5% =32 | VAF ≤ 2.5% =21 | Negative=95 | 0.717 |
| 3 mo | 3(0-9)% | 5(0-14)% | 5(1-10)% | 0.858 |
| 6 mo | 16(3-28)% | 19(2-36)% | 11(4-17)% | 0.561 |
| 1 yr | 22(7-36)% | 24(5-43)% | 17(9-24)% | 0.706 |
| 2 yr | 26(10-42)% | 24(5-43)% | 20(11-28)% | 0.743 |
| 3 yr | 26(10-42)% | 24(5-43)% | 28(16-40)% | 0.935 |
| RFS | 0<VAF<2.5% =32 | VAF ≤ 2.5% =21 | Negative=95 | 0.624 |
| 3 mo | 94(77-98)% | 90(67-98)% | 93(85-96)% | 0.913 |
| 6 mo | 75(56-87)% | 71(47-86)% | 84(75-90)% | 0.319 |
| 1 yr | 66(47-79)% | 67(43-83)% | 74(64-81)% | 0.626 |
| 2 yr | 61(42-76)% | 67(43-83)% | 69(59-78)% | 0.722 |
| 3 yr | 61(42-76)% | 56(27-77)% | 61(48-72)% | 0.926 |
| OS | 0<VAF<2.5% =32 | VAF ≤ 2.5% =21 | Negative=95 | 0.642 |
| 3 mo | 97(80-100)% | 95(71-99)% | 98(92-99)% | 0.84 |
| 6 mo | 84(66-93)% | 90(67-98)% | 92(84-96)% | 0.591 |
| 1 yr | 75(56-87)% | 86(62-95)% | 84(75-90)% | 0.516 |
| 2 yr | 66(46-80)% | 73(46-88)% | 75(65-83)% | 0.655 |
| 3 yr | 66(46-80)% | 61(29-82)% | 67(54-78)% | 0.918 |

**Supplemental Figure 5. NGS MRD status for *IDH1*-mutated AML patients and the association with clinical outcomes after allogeneic hematopoietic cell transplant stratified by baseline mutation groups.** Cumulative incidence of non-relapse mortality (NRM, top left) and relapse (top right), relapse-free survival (RFS, bottom left) and overall survival (OS, bottom right) shown at 24 months based on baseline patient mutation groups (A) *IDH1*-mutated AML patients with either *NPM1* and/or *FLT3*-ITD mutations at baseline (n=69) and (B) *IDH1*-mutated AML patients without *NPM1* or *FLT3*-ITD mutations at baseline (n=79). Point estimates at different time points are shown in the table (far right). Overall P values: Gray's test for non-relapse mortality (NRM) and relapse; log-rank test for relapse-free survival (RFS) and overall survival (OS). P values for pointwise estimations at different time points: z-test. Confidence interval, CI; Probability, prob; Month, mo; Year, yr; pos, positive; neg, negative.

**A**

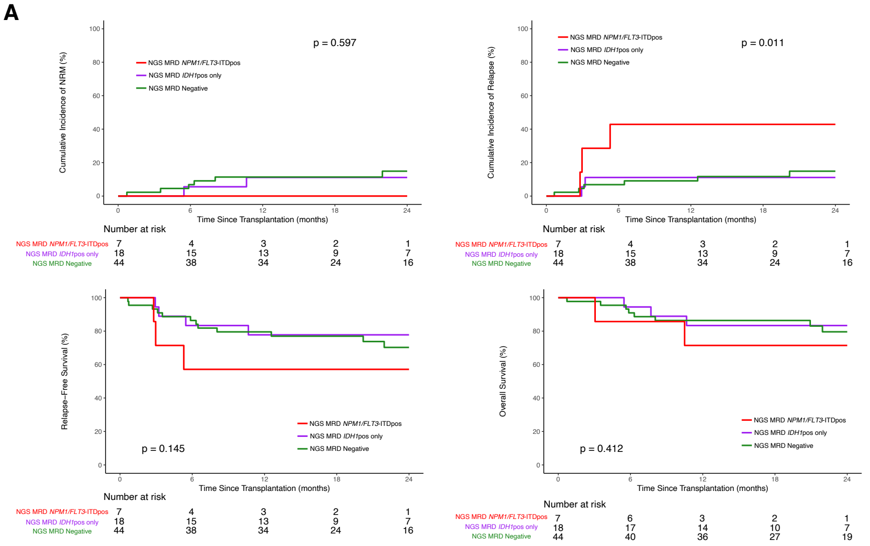


***NPM1/FLT3*-ITD Baseline Patients NGS MRD Status**

| Outcomes | Prob (95% CI) | Prob (95% CI) | Prob (95% CI) | p-value |
|---|---|---|---|---|
| NRM | *NPM1/FLT3*-ITDpos=7 | *IDH1*pos only=18 | MRD Negative=44 | 0.597 |
| 3 mo | 0(0-0)% | 0(0-0)% | 2(0-7)% | NA |
| 6 mo | 0(0-0)% | 6(0-16)% | 7(0-14)% | 0.127 |
| 1 yr | 0(0-0)% | 11(0-26)% | 11(2-21)% | 0.022 |
| 2 yr | 0(0-0)% | 11(0-26)% | 15(3-26)% | 0.014 |
| Relapse | *NPM1/FLT3*-ITDpos=7 | *IDH1*pos only=18 | MRD Negative=44 | 0.011 |
| 3 mo | 29(0-65)% | 6(0-16)% | 5(0-11)% | 0.444 |
| 6 mo | 43(2-83)% | 11(0-26)% | 7(0-14)% | 0.214 |
| 1 yr | 43(2-83)% | 11(0-26)% | 9(0-18)% | 0.277 |
| 2 yr | 43(2-83)% | 11(0-26)% | 15(4-26)% | 0.353 |
| RFS | *NPM1/FLT3*-ITDpos=7 | *IDH1*pos only=18 | MRD Negative=44 | 0.145 |
| 3 mo | 71(26-92)% | 94(67-99)% | 93(80-98)% | 0.434 |
| 6 mo | 57(17-84)% | 83(57-94)% | 86(72-94)% | 0.32 |
| 1 yr | 57(17-84)% | 78(51-91)% | 80(64-89)% | 0.522 |
| 2 yr | 57(17-84)% | 78(51-91)% | 70(53-82)% | 0.598 |
| OS | *NPM1/FLT3*-ITDpos=7 | *IDH1*pos only=18 | MRD Negative=44 | 0.412 |
| 3 mo | 100% | 100% | 98(85-100)% | NA |
| 6 mo | 86(33-98)% | 94(67-99)% | 91(78-96)% | 0.781 |
| 1 yr | 71(26-92)% | 83(57-94)% | 86(72-94)% | 0.694 |
| 2 yr | 71(26-92)% | 83(57-94)% | 80(63-89)% | 0.819 |

**B**

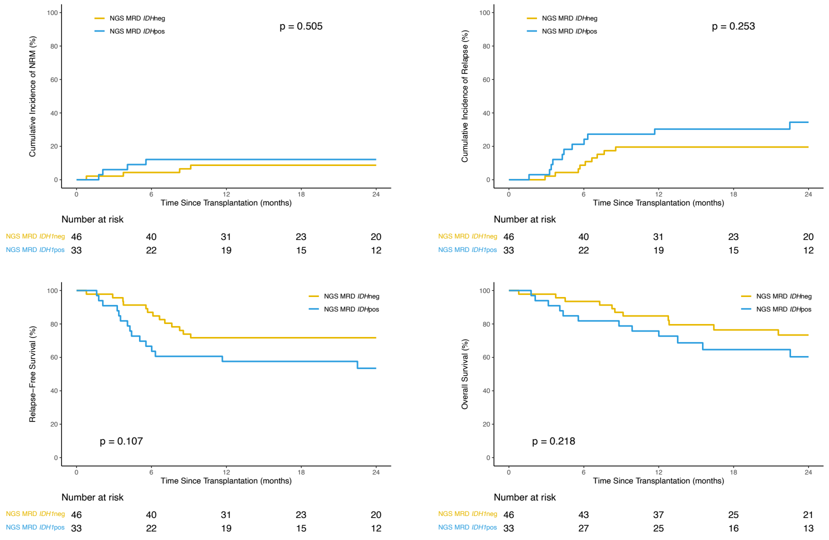


***IDH1* Only Baseline Patients (without *NPM1* or *FLT3*-ITD) NGS MRD Status**

| Outcomes | Prob (95% CI) | Prob (95% CI) | p-value |
|---|---|---|---|
| NRM | NGS MRD *IDH*neg=46 | NGS MRD *IDH*pos=33 | 0.505 |
| 3 mo | 2(0-6)% | 6(0-14)% | 0.413 |
| 6 mo | 4(0-10)% | 12(1-23)% | 0.234 |
| 1 yr | 9(0-17)% | 12(1-23)% | 0.632 |
| 2 yr | 9(0-17)% | 12(1-23)% | 0.632 |
| 3 yr | 9(0-17)% | 17(3-31)% | 0.34 |
| Relapse | NGS MRD *IDH*neg=46 | NGS MRD *IDH*pos=33 | 0.253 |
| 3 mo | 2(0-6)% | 3(0-9)% | 0.818 |
| 6 mo | 9(0-17)% | 21(7-35)% | 0.135 |
| 1 yr | 20(8-31)% | 30(14-46)% | 0.287 |
| 2 yr | 20(8-31)% | 34(17-52)% | 0.159 |
| 3 yr | 29(13-46)% | 34(17-52)% | 0.662 |
| RFS | NGS MRD *IDH*neg=46 | NGS MRD *IDH*pos=33 | 0.107 |
| 3 mo | 96(84-99)% | 91(74-97)% | 0.417 |
| 6 mo | 87(73-94)% | 67(48-80)% | 0.034 |
| 1 yr | 72(56-82)% | 58(39-72)% | 0.192 |
| 2 yr | 72(56-82)% | 53(35-69)% | 0.1 |
| 3 yr | 62(43-76)% | 49(30-65)% | 0.295 |
| OS | NGS MRD *IDH*neg=46 | NGS MRD *IDH*pos=33 | 0.218 |
| 3 mo | 98(86-100)% | 94(78-98)% | 0.406 |
| 6 mo | 93(81-98)% | 82(64-91)% | 0.127 |
| 1 yr | 85(71-92)% | 76(57-87)% | 0.324 |
| 2 yr | 73(57-84)% | 60(40-76)% | 0.259 |
| 3 yr | 68(50-81)% | 56(35-72)% | 0.307 |

**Supplemental Figure 6. NGS MRD status for *IDH1*-mutated AML patients and the association with clinical outcomes after allogeneic hematopoietic cell transplant stratified by conditioning intensity.** Cumulative incidence of non-relapse mortality (NRM, top left) and relapse (top right), relapse-free survival (RFS, bottom left) and overall survival (OS, bottom right) shown at 36 months based on NGS MRD *IDH1* status and conditioning intensity. Point estimates at different time points are shown in the table (far right). Overall P values: Gray's test for non-relapse mortality (NRM) and relapse; log-rank test for relapse-free survival (RFS) and overall survival (OS). P values for pointwise estimations at different time points: z-test. Confidence interval, CI; Probability, prob; Month, mo; Year, yr; pos, positive; neg, negative.

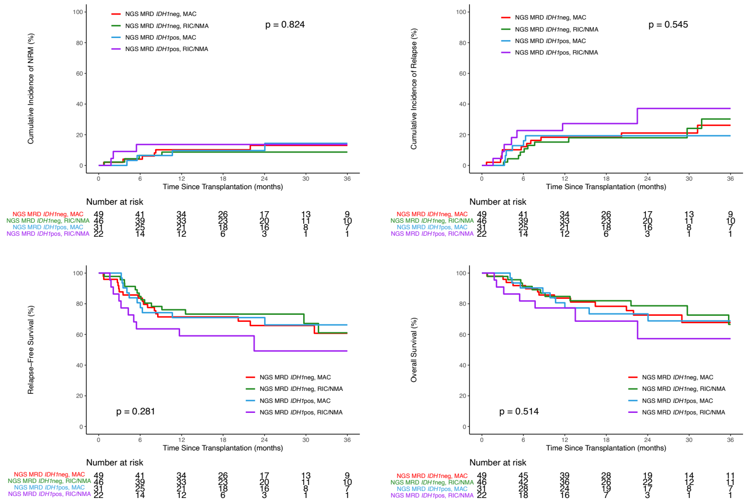


**NGS MRD *IDH1* Status and Conditioning Intensity**

| Outcomes | Prob (95% CI) | Prob (95% CI) | Prob (95% CI) | Prob (95% CI) | p-value |
|---|---|---|---|---|---|
| NRM | NGS MRD *IDH1*neg, MAC, n=49 | NGS MRD *IDH1*neg, RIC/NMA, n=46 | NGS MRD *IDH1*pos, MAC, n=31 | NGS MRD *IDH1*pos, RIC/NMA, n=22 | 0.824 |
| 3 mo | 2(0-6)% | 2(0-6)% | 0(0-0)% | 9(0-21)% | 0.252 |
| 6 mo | 4(0-10)% | 7(0-14)% | 6(0-15)% | 14(0-28)% | 0.681 |
| 1 yr | 10(2-19)% | 9(0-17)% | 10(0-20)% | 14(0-28)% | 0.953 |
| 2 yr | 13(3-23)% | 9(0-17)% | 10(0-20)% | 14(0-28)% | 0.89 |
| 3 yr | 13(3-23)% | 9(0-17)% | 14(1-28)% | 14(0-28)% | 0.852 |
| Relapse | NGS MRD *IDH1*neg, MAC, n=49 | NGS MRD *IDH1*neg, RIC/NMA, n=46 | NGS MRD *IDH1*pos, MAC, n=31 | NGS MRD *IDH1*pos, RIC/NMA, n=22 | 0.545 |
| 3 mo | 10(2-19)% | 0(0-0)% | 0(0-0)% | 9(0-21)% | NA |
| 6 mo | 12(3-22)% | 9(0-17)% | 13(1-25)% | 23(5-41)% | 0.573 |
| 1 yr | 18(7-29)% | 15(5-26)% | 19(5-34)% | 27(8-46)% | 0.755 |
| 2 yr | 21(9-33)% | 18(6-30)% | 19(5-34)% | 37(12-63)% | 0.609 |
| 3 yr | 26(11-41)% | 30(11-49)% | 19(5-34)% | 37(12-63)% | 0.623 |
| RFS | NGS MRD *IDH1*neg, MAC, n=49 | NGS MRD *IDH1*neg, RIC/NMA, n=46 | NGS MRD *IDH1*pos, MAC, n=31 | NGS MRD *IDH1*pos, RIC/NMA, n=22 | 0.281 |
| 3 mo | 88(75-94)% | 98(86-100)% | 100% | 82(59-93)% | <0.001 |
| 6 mo | 84(70-91)% | 85(71-92)% | 81(62-91)% | 64(40-80)% | 0.308 |
| 1 yr | 71(57-82)% | 76(61-86)% | 71(52-84)% | 59(36-76)% | 0.586 |
| 2 yr | 66(50-78)% | 73(58-84)% | 71(52-84)% | 49(24-70)% | 0.377 |
| 3 yr | 61(43-75)% | 61(40-77)% | 66(46-80)% | 49(24-70)% | 0.747 |
| OS | NGS MRD *IDH1*neg, MAC, n=49 | NGS MRD *IDH1*neg, RIC/NMA, n=46 | NGS MRD *IDH1*pos, MAC, n=31 | NGS MRD *IDH1*pos, RIC/NMA, n=22 | 0.514 |
| 3 mo | 98(86-100)% | 98(86-100)% | 100% | 91(68-98)% | <0.001 |
| 6 mo | 92(80-97)% | 91(78-97)% | 90(73-97)% | 82(59-93)% | 0.736 |
| 1 yr | 84(70-91)% | 85(71-92)% | 81(62-91)% | 77(54-90)% | 0.887 |
| 2 yr | 73(56-84)% | 79(63-88)% | 73(54-86)% | 57(27-79)% | 0.576 |
| 3 yr | 68(50-81)% | 67(44-82)% | 69(48-83)% | 57(27-79)% | 0.913 |

**Supplemental Table 1.** ***IDH1*-mutated AML patient baseline clinical characteristics.**

| Variable | Number of Patients (%) | Variable | Number of Patients (%) |
|---|---|---|---|
| **Age** | | **HCT-comorbidity index** | |
| <60 | 73 (49%) | 0 | 35 (24%) |
| ≥60 | 75 (51%) | 1-2 | 46 (31%) |
| **Sex** | | >2 | 67 (45%) |
| Female | 79 (53%) | **Karnofsky Score** | |
| Male | 69 (47%) | <90 | 68 (46%) |
| **Conditioning Intensity** | | ≥90 | 79 (53%) |
| MAC | 80 (54%) | **ATG usage** | |
| RIC/NMA | 68 (46%) | No | 124 (84%) |
| **Site-reported Flow Cytometry** | | Yes | 24 (16%) |
| Negative | 133 (90%) | **Race** | |
| Positive | 11 (7%) | Caucasian | 125 (84%) |
| **Graft Type** | | Other | 19 (13%) |
| Peripheral Blood | 111 (75%) | **ELN Risk Group** | |
| Cord Blood | 15 (10%) | Favorable | 30 (20%) |
| Bone Marrow | 22 (15%) | Intermediate | 68 (46%) |
| **Donor Type** | | Adverse | 50 (34%) |
| HLA-identical Sibling | 18 (12%) | **AML Group** | |
| Matched Unrelated | 96 (65%) | *De novo* | 122 (82%) |
| Haploidentical Related | 10 (7%) | Therapy-related | 9 (6%) |
| Cord Blood | 12 (8%) | Transformed MDS/MPN | 17 (11%) |
| Mismatched | 9 (6%) | **Baseline Mutation** | |
| Multiple Donors | 3 (2%) | *NPM1* and/or *FLT3*-ITD | 69 (47%) |

MAC, myeloablative; RIC, reduced intensity; NMA, nonmyeloablative; AML, acute myeloid leukemia

### Supplemental Table 2. Variants detected by next-generation sequencing in the blood of *IDH1* mutated AML patients prior to transplant conditioning.

| Subject ID | Gene | HGVSp | HGVSc | VAF | Alternate Observations | Deep Alternate Observations | Unique Alternate Observations | Depth | Deep Depth | Variant Consequence | Chromosome | Position | Reference | Alternate | Previously reported* | ddPCR validated | ddPCR VAF |
|---|---|---|---|---|---|---|---|---|---|---|---|---|---|---|---|---|---|
| CH 012119 | NPM1 | NP_002511.1:p.Trp288CysfsTer12 | NM_002520.6:c.860_863dup | 0.0001 | 4 | 1 | 1 | 37243 | 10054 | frameshift | 5 | 170837543 | C | CTCTG | Yes | Yes | 0.00015 |
| CH 012020 | IDH1 | NP_005887.2:p.Arg132Cys | NM_005896.3:c.394C>T | 0.006 | 280 | 118 | 117 | 36952 | 19630 | missense | 2 | 209113113 | G | A | Yes | Yes | 0.0087184 |
| CH 012505 | IDH1 | NP_005887.2:p.Arg132Ser | NM_005896.3:c.394C>A | 0.2104 | 6529 | 3600 | 562 | 30474 | 17113 | missense | 2 | 209113113 | G | T | Yes | Yes | 0.21690 |
| CH 012247 | IDH1 | NP_005887.2:p.Arg132His | NM_005896.3:c.395G>A | 0.0124 | 572 | 170 | 207 | 40301 | 13742 | missense | 2 | 209113112 | C | T | Yes | Yes | 0.01460 |
| CH 012139 | IDH1 | NP_005887.2:p.Arg132Cys | NM_005896.3:c.394C>T | 0.0142 | 496 | 203 | 179 | 28504 | 14249 | missense | 2 | 209113113 | G | A | Yes | Yes | 0.01799 |
| CH 012442 | IDH1 | NP_005887.2:p.Arg132His | NM_005896.3:c.395G>A | 0.0082 | 438 | 185 | 168 | 50814 | 22624 | missense | 2 | 209113112 | C | T | Yes | Yes | 0.00942 |
| CH 012476 | IDH1 | NP_005887.2:p.Arg132Gly | NM_005896.3:c.394C>G | 0.3379 | 12299 | 3240 | 700 | 35090 | 9589 | missense | 2 | 209113113 | G | C | Yes | Yes | 0.34389 |
| CH 012090 | IDH2 | NP_002159.2:p.Arg140Gln | NM_002168.3:c.419G>A | 0.0016 | 30 | 16 | 22 | 15073 | 9969 | missense | 15 | 90631934 | C | T | Yes | Yes | 0.00111 |
| CH 012177 | IDH1 | NP_005887.2:p.Arg132Cys | NM_005896.3:c.394C>T | 0.0627 | 2896 | 1236 | 429 | 40875 | 19728 | missense | 2 | 209113113 | G | A | Yes | Yes | 0.06833 |
| CH 012228 | IDH1 | NP_005887.2:p.Arg132Cys | NM_005896.3:c.394C>T | 0.0042 | 187 | 74 | 83 | 34772 | 17459 | missense | 2 | 209113113 | G | A | Yes | Yes | 0.00407 |
| CH 016941 | FLT3 | NP_004110.2:p.Glu598_Tyr599insPheAspPheArgGlu TyrGlu | NM_004119.2:c.1775_1795dup | 0.0002 | 17 | | 16 | 74735 | N/A | FLT3-ITD | 13 | 28608260 | T | TATTCATATTCTCTGAAATCAA | Yes | Not Done | Not Done |
| CH 016498 | IDH1 | NP_005887.2:p.Arg132Ser | NM_005896.3:c.394C>A | 0.0011 | 65 | 23 | 42 | 44701 | 21453 | missense | 2 | 209113113 | G | T | Yes | Yes | 0.00090 |
| CH 016870 | IDH1 | NP_005887.2:p.Arg132Gly | NM_005896.3:c.394C>G | 0.0106 | 298 | 166 | 133 | 26660 | 15691 | missense | 2 | 209113113 | G | C | Yes | Yes | 0.01208 |
| CH 016683 | IDH1 | NP_005887.2:p.Arg132Ser | NM_005896.3:c.394C>A | 0.0103 | 239 | 147 | 118 | 23713 | 14224 | missense | 2 | 209113113 | G | T | Yes | Yes | 0.00980 |
| CH 016673 | IDH1 | NP_005887.2:p.Arg132Gly | NM_005896.3:c.394C>G | 0.0195 | 600 | 345 | 221 | 30821 | 17734 | missense | 2 | 209113113 | G | C | Yes | Yes | 0.02029 |
| CH 016649 | IDH1 | NP_005887.2:p.Arg132Cys | NM_005896.3:c.394C>T | 0.0223 | 619 | 342 | 220 | 25060 | 15306 | missense | 2 | 209113113 | G | A | Yes | Yes | 0.02783 |
| CH 016764 | IDH1 | NP_005887.2:p.Arg132His | NM_005896.3:c.395G>A | 0.0022 | 83 | 37 | 52 | 28437 | 17182 | missense | 2 | 209113112 | C | T | Yes | Yes | 0.00320 |
| CH 016764 | IDH2 | NP_002159.2:p.Arg140Gln | NM_002168.3:c.419G>A | 0.4032 | 6319 | 4380 | 463 | 15760 | 10862 | missense | 15 | 90631934 | C | T | Yes | Yes | 0.39821 |
| CH 016546 | FLT3 | NP_004110.2:p.Arg595_Leu601dup | NM_004119.2:c.1784_1804dup | 0.0004 | 30 | | 17 | 84776 | N/A | FLT3-ITD | 13 | 28608251 | T | TTGAGATCATATTCATATTCTC | Yes | Not Done | Not Done |
| CH 016546 | NPM1 | NP_002511.1:p.Trp288CysfsTer12 | NM_002520.6:c.860_863dup | 0.0009 | 31 | 4 | 20 | 33577 | 10783 | frameshift | 5 | 170837543 | C | CTCTG | Yes | Yes | 0.00087 |
| CH 016548 | NPM1 | NP_002511.1:p.Trp288CysfsTer12 | NM_002520.6:c.860_863dup | 0.0001 | 3 | 0 | 1 | 22233 | 8099 | frameshift | 5 | 170837543 | C | CTCTG | Yes | Yes | 0.00023 |
| CH 016989 | IDH1 | NP_005887.2:p.Arg132Cys | NM_005896.3:c.394C>T | 0.0292 | 1011 | 575 | 294 | 32448 | 19666 | missense | 2 | 209113113 | G | A | Yes | Yes | 0.03584 |
| CH 016748 | IDH1 | NP_005887.2:p.Arg132Cys | NM_005896.3:c.394C>T | 0.0355 | 1232 | 729 | 313 | 32558 | 20552 | missense | 2 | 209113113 | G | A | Yes | Yes | 0.03998 |
| CH 016748 | NPM1 | NP_002511.1:p.Trp288CysfsTer12 | NM_002520.6:c.860_863dup | 0.0005 | 17 | 3 | 10 | 34071 | 15101 | frameshift | 5 | 170837543 | C | CTCTG | Yes | Yes | 0.00019 |
| CH 016755 | IDH2 | NP_002159.2:p.Arg140Gln | NM_002168.3:c.419G>A | 0.0192 | 244 | 167 | 105 | 13044 | 8692 | missense | 15 | 90631934 | C | T | Yes | Yes | 0.01828 |
| CH 016486 | IDH1 | NP_005887.2:p.Arg132Cys | NM_005896.3:c.394C>T | 0.0021 | 46 | 28 | 35 | 20823 | 13423 | missense | 2 | 209113113 | G | A | Yes | Yes | 0.00329 |
| CH 016466 | IDH1 | NP_005887.2:p.Arg132Cys | NM_005896.3:c.394C>T | 0.0026 | 84 | 54 | 54 | 32440 | 20414 | missense | 2 | 209113113 | G | A | Yes | Yes | 0.00252 |
| CH 016799 | IDH1 | NP_005887.2:p.Arg132Gly | NM_005896.3:c.394C>G | 0.0019 | 54 | 25 | 41 | 23572 | 13146 | missense | 2 | 209113113 | G | C | Yes | Yes | 0.00150 |
| CH 017017 | IDH1 | NP_005887.2:p.Arg132Cys | NM_005896.3:c.394C>T | 0.0022 | 62 | 35 | 45 | 25384 | 15728 | missense | 2 | 209113113 | G | A | Yes | Yes | 0.00263 |
| CH 016447 | IDH1 | NP_005887.2:p.Arg132Cys | NM_005896.3:c.394C>T | 0.0268 | 1085 | 608 | 293 | 37934 | 22717 | missense | 2 | 209113113 | G | A | Yes | Yes | 0.02671 |
| CH 016484 | IDH1 | NP_005887.2:p.Arg132Cys | NM_005896.3:c.394C>T | 0.0056 | 203 | 117 | 97 | 33383 | 20940 | missense | 2 | 209113113 | G | A | Yes | Yes | 0.00610 |
| CH 016589 | IDH1 | NP_005887.2:p.Arg132Cys | NM_005896.3:c.394C>T | 0.0025 | 105 | 50 | 60 | 32822 | 19978 | missense | 2 | 209113113 | G | A | Yes | Yes | 0.00343 |
| CH 016650 | IDH2 | NP_002159.2:p.Arg140Gln | NM_002168.3:c.419G>A | 0.3236 | 6062 | 4134 | 472 | 18676 | 12774 | missense | 15 | 90631934 | C | T | Yes | Yes | 0.31988 |
| CH 016995 | IDH1 | NP_005887.2:p.Arg132Cys | NM_005896.3:c.394C>T | 0.0272 | 904 | 536 | 236 | 32960 | 19677 | missense | 2 | 209113113 | G | A | Yes | Yes | 0.02704 |
| CH 016829 | IDH1 | NP_005887.2:p.Arg132Ser | NM_005896.3:c.394C>A | 0.0011 | 124 | 20 | 76 | 51081 | 17490 | missense | 2 | 209113113 | G | T | Yes | Yes | 0.00229 |
| CH 016825 | IDH1 | NP_005887.2:p.Arg132His | NM_005896.3:c.395G>A | 0.3064 | 14168 | 7169 | 872 | 44813 | 23394 | missense | 2 | 209113112 | C | T | Yes | Yes | 0.31749 |
| CH 016582 | IDH2 | NP_002159.2:p.Arg140Gln | NM_002168.3:c.419G>A | 0.001 | 58 | 19 | 35 | 31330 | 19911 | missense | 15 | 90631934 | C | T | Yes | Yes | 0.00131 |
| CH 016969 | IDH1 | NP_005887.2:p.Arg132Cys | NM_005896.3:c.394C>T | 0.0099 | 555 | 237 | 200 | 47267 | 23840 | missense | 2 | 209113113 | G | A | Yes | Yes | 0.01298 |
| CH 017043 | IDH1 | NP_005887.2:p.Arg132Cys | NM_005896.3:c.394C>T | 0.0066 | 275 | 135 | 136 | 36039 | 20601 | missense | 2 | 209113113 | G | A | Yes | Yes | 0.00789 |
| CH 017073 | IDH1 | NP_005887.2:p.Arg132Cys | NM_005896.3:c.394C>T | 0.0423 | 1356 | 764 | 330 | 30626 | 18074 | missense | 2 | 209113113 | G | A | Yes | Yes | 0.04679 |
| CH 016893 | IDH1 | NP_005887.2:p.Arg132Cys | NM_005896.3:c.394C>T | 0.0826 | 4273 | 1703 | 568 | 44684 | 20612 | missense | 2 | 209113113 | G | A | Yes | Yes | 0.10241 |
| CH 016946 | IDH1 | NP_005887.2:p.Arg132Cys | NM_005896.3:c.394C>T | 0.0336 | 1023 | 511 | 275 | 26841 | 15202 | missense | 2 | 209113113 | G | A | Yes | Yes | 0.03914 |
| CH 016958 | IDH1 | NP_005887.2:p.Arg132Gly | NM_005896.3:c.394C>G | 0.3531 | 6344 | 3132 | 534 | 18124 | 8870 | missense | 2 | 209113113 | G | C | Yes | Yes | 0.35364 |
| CH 016964 | IDH2 | NP_002159.2:p.Arg140Gln | NM_002168.3:c.419G>A | 0.0028 | 40 | 27 | 26 | 13565 | 9511 | missense | 15 | 90631934 | C | T | Yes | Not Done | Not Done |
| CH 017008 | IDH1 | NP_005887.2:p.Arg132His | NM_005896.3:c.395G>A | 0.0165 | 587 | 253 | 200 | 34690 | 15290 | missense | 2 | 209113112 | C | T | Yes | Yes | 0.01861 |
| CH 017078 | IDH1 | NP_005887.2:p.Arg132Cys | NM_005896.3:c.394C>T | 0.0024 | 120 | 42 | 56 | 33768 | 17434 | missense | 2 | 209113113 | G | A | Yes | Yes | 0.00373 |
| CH 017022 | IDH1 | NP_005887.2:p.Arg132His | NM_005896.3:c.395G>A | 0.4846 | 25024 | 7806 | 884 | 51843 | 16108 | missense | 2 | 209113112 | C | T | Yes | Yes | 0.48050 |
| CH 017053 | IDH1 | NP_005887.2:p.Arg132Gly | NM_005896.3:c.394C>G | 0.001 | 31 | 17 | 22 | 34789 | 16631 | missense | 2 | 209113113 | G | C | Yes | Yes | 0.00050 |
| CH 017091 | IDH1 | NP_005887.2:p.Arg132Gly | NM_005896.3:c.394C>G | 0.0121 | 479 | 224 | 158 | 35214 | 18509 | missense | 2 | 209113113 | G | C | Yes | Yes | 0.00598 |
| CH 017495 | IDH1 | NP_005887.2:p.Arg132Gly | NM_005896.3:c.394C>G | 0.1225 | 5616 | 1950 | 555 | 43459 | 15916 | missense | 2 | 209113113 | G | C | Yes | Yes | 0.12944 |
| CH 017385 | IDH1 | NP_005887.2:p.Arg132His | NM_005896.3:c.395G>A | 0.0224 | 1114 | 363 | 301 | 40319 | 16232 | missense | 2 | 209113112 | C | T | No | Yes | 0.031 |
| CH 017402 | IDH1 | NP_005887.2:p.Arg132His | NM_005896.3:c.395G>A | 0.0352 | 1349 | 445 | 353 | 34420 | 12628 | missense | 2 | 209113112 | C | T | No | Yes | 0.0424 |
| CH 017402 | NPM1 | NP_002511.1:p.Trp288CysfsTer12 | NM_002520.6:c.863_864insCTTG | 0.0386 | 1035 | 143 | 317 | 26807 | 3750 | frameshift | 5 | 170837545 | C | CTGCT | No | Not Done | Not Done |
| CH 017425 | FLT3 | NP_004110.2:p.Gly583_Leu601dup | NM_004119.2:c.1747_1803dup | 0.0118 | 576 | | 63 | 48885 | N/A | FLT3-ITD | 13 | 28608253 | G | GAGATCATATTCATATTCTCTGAAATCAACGTAGA AGTACTCATTATCTGAGGAGCCG | No | Not Done | Not Done |
| CH 017425 | NPM1 | NP_002511.1:p.Trp288CysfsTer12 | NM_002520.6:c.860_863dup | 0.0014 | 24 | 4 | 17 | 16781 | 2673 | frameshift | 5 | 170837543 | C | CTCTG | No | Not Done | Not Done |
| CH 017430 | IDH1 | NP_005887.2:p.Arg132Cys | NM_005896.3:c.393_394delinsCT | 0.0148 | 630 | 166 | 215 | 39016 | 11222 | missense | 2 | 209113113 | GA | AG | No | Not Done | Not Done |
| CH 017433 | IDH2 | NP_002159.2:p.Arg172Lys | NM_002168.3:c.515G>A | 0.004 | 121 | 70 | 66 | 27354 | 17655 | missense | 15 | 90631838 | C | T | No | Yes | 0.0042 |
| CH 017444 | IDH1 | NP_005887.2:p.Arg132Cys | NM_005896.3:c.394C>T | 0.0028 | 154 | 43 | 81 | 38875 | 15569 | missense | 2 | 209113113 | G | A | No | Yes | 0.0047 |
| CH 017474 | IDH1 | NP_005887.2:p.Arg132Cys | NM_005896.3:c.394C>T | 0.0147 | 481 | 105 | 158 | 33924 | 7148 | missense | 2 | 209113113 | G | A | No | Yes | 0.0143 |
| CH 017505 | IDH1 | NP_005887.2:p.Arg132Cys | NM_005896.3:c.394C>T | 0.0255 | 900 | 429 | 276 | 33139 | 16820 | missense | 2 | 209113113 | G | A | No | Yes | 0.0273 |
| CH 017549 | IDH1 | NP_005887.2:p.Arg132Cys | NM_005896.3:c.394C>T | 0.0064 | 195 | 59 | 97 | 25964 | 9216 | missense | 2 | 209113113 | G | A | No | Yes | 0.008 |
| CH 017573 | IDH2 | NP_002159.2:p.Arg140Gln | NM_002168.3:c.419G>A | 0.0046 | 90 | 33 | 57 | 16306 | 7124 | missense | 15 | 90631934 | C | T | No | Yes | 0.0058 |
| CH 017580 | IDH1 | NP_005887.2:p.Arg132Cys | NM_005896.3:c.394C>T | 0.0101 | 343 | 58 | 140 | 32296 | 5740 | missense | 2 | 209113113 | G | A | No | Yes | 0.0109 |
| CH 017589 | IDH1 | NP_005887.2:p.Arg132Cys | NM_005896.3:c.394C>T | 0.1193 | 3316 | 1242 | 485 | 26777 | 10414 | missense | 2 | 209113113 | G | A | No | Yes | 0.1257 |
| CH 017592 | IDH1 | NP_005887.2:p.Arg132Leu | NM_005896.3:c.395G>T | 0.1298 | 4953 | 1451 | 531 | 35798 | 11180 | missense | 2 | 209113112 | C | A | No | Not Done | Not Done |
| CH 017607 | IDH1 | NP_005887.2:p.Arg132Cys | NM_005896.3:c.394C>T | 0.0566 | 1518 | 904 | 354 | 25707 | 15960 | missense | 2 | 209113113 | G | A | No | Yes | 0.0586 |
| CH 017634 | IDH1 | NP_005887.2:p.Arg132His | NM_005896.3:c.395G>A | 0.4115 | 10535 | 5534 | 655 | 25471 | 13449 | missense | 2 | 209113112 | C | T | No | Yes | 0.4048 |
| CH 017640 | IDH1 | NP_005887.2:p.Arg132Cys | NM_005896.3:c.394C>T | 0.0094 | 466 | 132 | 183 | 39332 | 13980 | missense | 2 | 209113113 | G | A | No | Yes | 0.0121 |
| CH 017643 | IDH1 | NP_005887.2:p.Arg132Gly | NM_005896.3:c.394C>G | 0.0352 | 1572 | 462 | 384 | 42114 | 13117 | missense | 2 | 209113113 | G | C | No | Yes | 0.037 |
| CH 017644 | IDH2 | NP_002159.2:p.Arg140Gln | NM_002168.3:c.419G>A | 0.0012 | 36 | 15 | 22 | 18460 | 12657 | missense | 15 | 90631934 | C | T | No | Yes | 0.0018 |
| CH 017645 | IDH1 | NP_005887.2:p.Arg132His | NM_005896.3:c.395G>A | 0.001 | 79 | 17 | 53 | 40408 | 17558 | missense | 2 | 209113112 | C | T | No | Yes | 0.0018 |

VAF, variant allele frequency; ddPCR, digital droplet PCR; N/A, not applicable
*Variants previously reported in Dillon *et al*, JAMA, 2023. doi:10.1001/jama.2023.1363
